# Deep learning-based assessment of ulcerative colitis activity from full-length endoscopic videos with spatial characterisation and histological correlation

**DOI:** 10.64898/2026.09.16.26363201

**Authors:** Alex Bogush, Alexandros Toskas, George Ralli, Dylan Windell, Paul Aljabar, Mark DeLegee, Alissa Walsh, John P Thomas, Phil Wakefield, Caitlin Langford, Eve Fryer, Rob Goldin, Noriko Suzuki, Jon Landy

**Affiliations:** Perspectum Ltd, Oxford, UK; St Mark’s Hospital, London North West University Healthcare NHS Trust, London, UK; IQVIA Inc., Falls Church, VA, USA; Oxford University Hospitals NHS Foundation Trust Oxford, UK; Imperial College, London, UK

**Keywords:** Ulcerative colitis, Endoscopy, Deep learning, Artificial intelligence, Inflammation, Mayo Endoscopic Score, UCEIS, Continuous Disease Score, Video analysis, Disease activity, Histology

## Abstract

**Background and study aims:** Endoscopic assessment of ulcerative colitis (UC) is central to clinical decision-making however remains subjective and limited in characterisation of disease distribution. Artificial intelligence (AI) enables analysis of entire endoscopic examination, rather than relying on selected views. We aimed to develop and validate a deep learning model for automated assessment of UC severity from full-length endoscopic videos, introducing spatial representation of inflammation (Continuous Disease Score, CDS), and assess histological correlation.

**Patients and methods:** Full-length endoscopy videos from adult patients with UC undergoing colonoscopy or flexible sigmoidoscopy were analysed; isolated proctitis was excluded. Videos were segmented and annotated using Mayo Endoscopic Score (MES) and Ulcerative Colitis Endoscopic Index of Activity (UCEIS). A deep learning model was trained for frame-level quality control and severity prediction, enabling analysis of full-length videos. Performance was evaluated using quadratic weighted kappa (QWK) and Cohen’s kappa, with patient-level separation between datasets. CDS was derived from UCEIS predictions to quantify cumulative inflammatory burden, spatial extent of disease and histological prediction.

**Results:** A total of 67 videos from 59 patients were included. The model demonstrated agreement for remission classification (MES=0 κ 0.76; UCEIS≤1 κ 0.84). CDS enabled quantification of inflammatory burden and revealed spatial heterogeneity not reflected in categorical scores. Agreement with histology was strong (AUROC 0.84–0.87).

**Conclusions:** AI-based analysis enables automated assessment of UC activity from full-length endoscopic videos, including remission detection and estimation of histological healing. CDS provides continuous characterisation of inflammatory burden and disease extent beyond conventional categorical scores, with potential to support more standardised assessment in clinical trials and practice.

## Introduction

The incidence and prevalence of inflammatory bowel diseases are increasing worldwide [1]. Objective and reliable measurements are important in disease assessment and prognostication. Endoscopic healing is the preferred long-term treatment goal in ulcerative colitis (UC) [2]. Histologic remission may be an adjunct to endoscopic remission and is associated with long-term remission and cancer reduction [2]. Pancolonic histological evaluation is necessary for assessment of disease distribution and activity. However, this requires multiple biopsies, increasing cost and limiting timely clinical decision making.

There remain significant barriers to implementing endoscopic and histological healing as a treatment target. Endoscopic scoring systems such as the Mayo endoscopic subscore (MES) and the Ulcerative Colitis Endoscopic Index of Severity (UCEIS) have been established and validated [3, 4]. These are widely used in both clinical trials and routine practice. However, both MES and UCEIS are also subject to interobserver variability and these scoring systems do not provide an overview of the distribution or gradient of disease severity throughout the colon [3–5]. Interobserver variability is greater for less experienced endoscopists [3]. Furthermore, prior knowledge of clinical symptoms may affect threshold evaluation of endoscopic subscores such as bleeding [4].

To mitigate these limitations and improve interobserver variability, central reading has been used in UC clinical trials. However, this increases the time and financial costs of studies and does not enhance the quality of video data captured. This is not a solution for routine clinical practice where standardisation would improve clinical decision making.

Recent studies have suggested a role for machine learning in image analysis in the field of endoscopy. Encouraging results are emerging with the application of machine learning models in endoscopic and histological assessment for UC [6–14]. However, several previously reported AI systems required specialised imaging modalities or hardware-dependent techniques not routinely available in standard clinical practice, such as virtual chromoendoscopy or proprietary image enhancement platforms [9, 14]. Others have reported systems using still images or restricted to single anatomical colonic segments rather than full-length routine HD-WLE video.

In this work, we acquired a dataset consisting of full-length endoscopy videos in UC patients, along with paired histological reads from biopsies taken during the endoscopies. The primary objective was to develop machine learning models to assess endoscopic activity or remission from the full-length videos. Secondary objectives were to assess the ability of the model to grade disease severity, to generate a continuous disease score compared with MES and UCEIS, and to detect histological remission.

## Material and Methods

### Study Cohort

Full-length, unaltered, de-identified videos were recorded from UC patients undergoing endoscopic evaluation by sigmoidoscopy or colonoscopy. Biopsy samples were taken during the endoscopy exams according to routine clinical practice. Patients were recruited between 2022 and 2023 from two sites of the West Hertfordshire Hospitals NHS Trust (NCT05000242) [15]. Exclusion criteria included contraindication to endoscopic procedure or biopsies, and patients with disease limited to the rectum. The study was approved by the London - Queen Square Research Ethics Committee (REC: 20/LO/0349). All patients gave informed consent to participate in the study. The study was conducted in accordance with the Declaration of Helsinki (2013) and Good Clinical Practice guidelines.

A total of 67 full-length endoscopic videos were collected from 59 patients with UC. Baseline demographic and clinical characteristics are summarised in Table 1. Videos included examinations of multiple anatomical segments per patient and were analysed at both segment and whole-colon levels. Procedures were undertaken using 290 or 1500 series colonoscopes (Olympus, Japan). All procedures were performed with high-definition white light endoscopy (HD-WLE). An overview of the AI pipeline for HD-WLE video analysis is shown in Figure 1A.

**Table 1.** Study populations. *Shown as n (%). ^#^Shown as Mean± SD.

| **Demographics<sup>#</sup>** |  |
| --- | --- |
| Age (yr) | 43 ±15 |
| Male | 32(54%) |
| Female | 27(46%) |
| BMI | 25.3 ±6.9 |
| **Non-invasive tests<sup>#</sup>** |  |
| Haemoglobin | 134 ±19 (n=42) |
| C-Reactive Protein | 12.6 ±15.9 (n=34) |
| Faecal Calprotectin | 1391 ±1600 (n=16) |
| Albumin | 37.0 ±5.1 (n=40) |
| **Treatment\*** |  |
| Topical (5-ASA) | 33 (56%) |
| Biologic/Small Molecule<br>(TNF-α, Ustekinumab,<br>Vedolizumab, Tofacitinib) | 20 (34%) |
| Steroid | 1 (2%) |

**Figure 1A.**
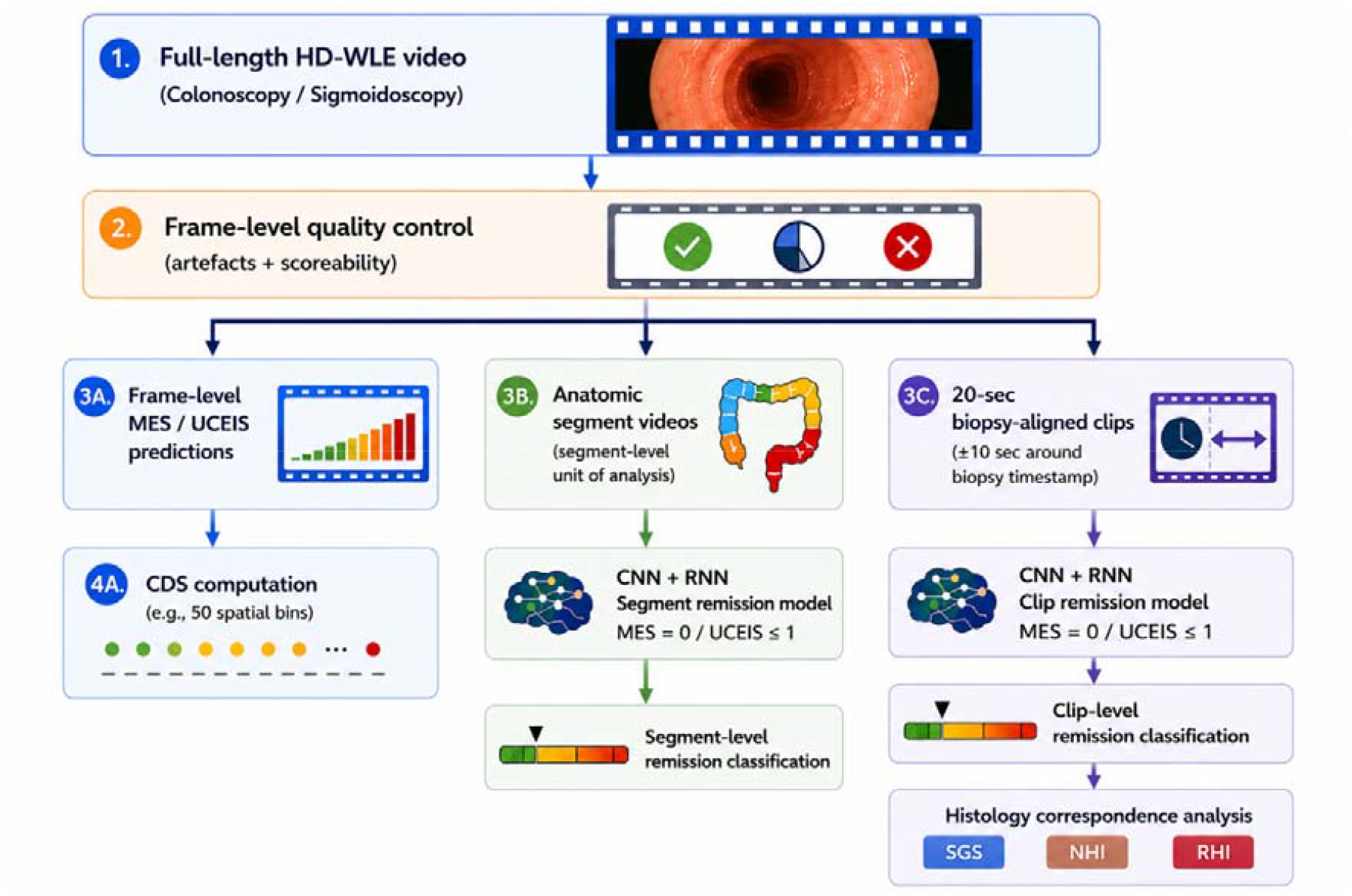
AI pipeline architecture for analysis of full-length HD-WLE videos

**Figure 1B.**
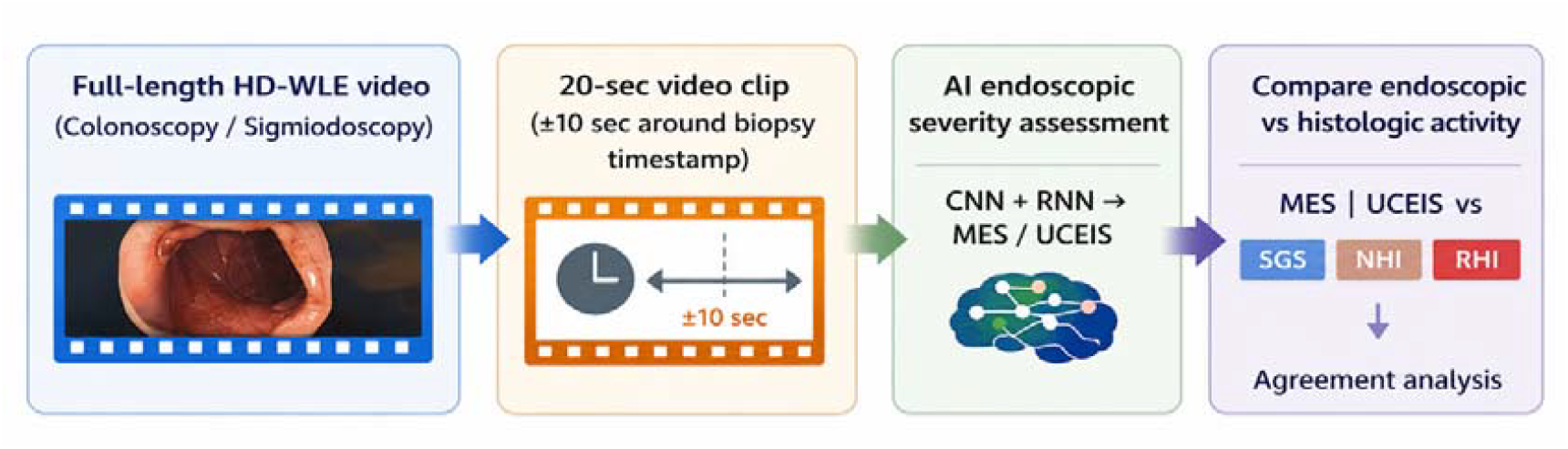
Biopsy-aligned endoscopy and histology remission analysis

### Dataset extraction and annotation

A total of 2168 still frames were extracted from the endoscopic videos. These frames were assigned Mayo Endoscopic Score (MES) and Ulcerative Colitis Endoscopic Index of Severity (UCEIS) scores by three endoscopists (NS, JL, AT). 1288 frames were deemed un-scoreable by the endoscopists due to poor quality and excluded from further analysis.

Artefact load included factors such as illumination, blur, debris, reflections, instruments, post biopsy bleeding, and imaging modes such as NBI. For the remaining 880 frames, the MES and UCEIS scores from the three endoscopists were combined into a single consensus score by majority vote, with the most senior endoscopist (NS) acting as a tie-breaker where all three scores were different.

To assess endoscopic severity, videos were split on anatomic segments (rectum, descending, transverse, and ascending colon) using timestamps provided by one endoscopist (JL). A total of 151 anatomically defined videos were extracted. These video segments were assigned labels of MES and UCEIS scores from two endoscopists (JL, AT). Discrepant cases were reviewed at the consensus meeting where disagreement was resolved.

To facilitate comparisons between endoscopic and histological scores, for each biopsy event in the video data, a separate 20-second video clip centred on the biopsy timestamp (±10 seconds) was extracted per biopsies. A total of 316 clips were extracted. These clips were assigned labels of MES and UCEIS scores from two endoscopists (JL, AT). Discrepant cases were reviewed at a consensus meeting where disagreement was resolved. An overview of the AI pipeline for biopsy-aligned clips is shown in Figure 1B.

Biopsy samples were fixed in formalin, stained with haematoxylin and eosin and digitally scanned using whole-slide scanners. Biopsies were independently scored by two expert IBD histopathologists (RG/EF), blinded to endoscopic and clinical data, using a scoring template designed to translate into the Simplified Geboes Score (SGS), the Nancy Histological Index (NHI), and the Robarts Histopathology Index (RHI). Histological remission was defined as SGS≤2A.1, NHI=0 or RHI≤3 [16]. Agreement analyses were performed against both pathologists; agreement between pathologists was substantial to almost perfect for SGS, NHI and RHI (QWK 0.87, 0.77 and 0.90, respectively), therefore only comparisons with the senior histopathologist (RG) are presented.

### Frame-level scoring models

For the frame level dataset, a series of ResNet152 models were trained to predict individual MES and UCEIS scores, endoscopic remission defined as MES=0, and UCEIS≤1, UCEIS severity grading using four categories (0, 1–3, 4–5, and 6–8). To train the models, the frame-level dataset was split at the patient level into 36 patients (527 frames) for training, 11 patients (181 frames) for validation and 11 patients (172 frames) as held-out test data. Label distributions for each of these datasets are given in Supplementary Table 1.

A further ResNet152 model was trained to predict whether a given frame was of sufficient quality to be scored or not, based on a subset of the frames assessed by the three endoscopists. This dataset consisted of 2168 frames, of which 880 were marked as scoreable. Individual artefacts were not classified separately, but instead the model was trained to predict whether a frame was either scoreable or not.

All ResNet152 models were initialised based on pre-trained weights from the ImageNet database.

### Continuous disease scoring (CDS)

A CDS approach was adapted from the cumulative disease scoring framework [12] to provide a quantitative measure of inflammatory burden across the colon by integrating disease severity and extend and distribution. We extended this framework to perform segment-level and whole-colon analysis using MES and UCEIS predictions.

For CDS computation, each scorable video frame within an anatomical segment was assigned an ordinal severity value corresponding to the predicted MES or UCEIS category. Anatomical segments were divided into a fixed number of bins (50 bins per segment) spanning the full duration of the segment. For each bin, the proportion of scorable frames was determined using the scorable-frame model. Bins with more than 15% scorable frames were included in the analysis. Within each included bin, all frames were scored, and the bin severity was defined by the highest severity score observed among its frames.

Segment-level CDS values were calculated by summing bin-level severity contributions and weighting them according to the proportion of included bins, enabling comparison across anatomical regions and between patients. Whole-colon CDS values were derived by combining normalised CDS values from individual colonic segments (right colon, transverse colon, left colon, and rectum), yielding a single continuous metric representing global inflammatory burden.

### Segment-level remission analysis

To analyse anatomically defined video segments, a spatio-temporal deep learning architecture (segment remission model) was developed to model both spatial and temporal patterns within each segment. Only frames classified as scorable by the quality-control model described above were included in the analysis. Individual frames were processed using a ResNet152 convolutional neural network to generate 2048-dimensional feature representations. These frame-level features were subsequently analysed using a recurrent neural network (RNN) with long short-term memory (LSTM) units, producing a single prediction for the entire anatomical segment.

This architecture was trained to predict endoscopic remission as defined by MES=0 and UCEIS≤1. To train the models, the segment-level dataset was split at the patient level into 34 patients (81 segments) for training, 8 patients (20 segments) for validation, and 17 patients (50 segments) as held-out test data. Label distributions for each dataset are provided in Supplementary Table 2.

### Biopsy-aligned clip-level analysis and correspondence with histology

To analyse biopsy-aligned video clips, the same spatio-temporal deep learning architecture (clip remission model) was applied to model both spatial and temporal patterns within each clip. Only frames classified as scorable by the quality-control model were included in the analysis.

This architecture was trained to predict endoscopic remission as defined by MES=0 and UCEIS≤1. To train the models, the clip-level dataset was split at the patient level into 27 patients (160 clips) for training, 12 patients (56 clips) for validation, and 20 patients (100 clips) as held-out test data. Label distributions for each dataset are provided in Supplementary Table 3.

### Statistical analysis

Model performance was assessed using contingency tables, with results reported as accuracy, sensitivity, specificity, positive predictive value (PPV), negative predictive value (NPV), and area under the receiver operating characteristic curve (AUROC). 95% confidence intervals were calculated using non-parametric bootstrapping with 1,000 resamples.

Agreement between AI predictions and expert reference standards was assessed using Cohen’s kappa coefficient and quadratic weighted kappa, interpreted according to established thresholds (≤0, no agreement; 0.01–0.20, slight; 0.21–0.40, fair; 0.41–0.60, moderate; 0.61–0.80, substantial; 0.81–1.00, almost perfect agreement).

The study was conducted and reported in accordance with the Checklist for Artificial Intelligence in Medical Imaging (CLAIM) [17] and the TRIPOD guidelines [18] for prediction model development and validation (Supplementary Tables 4-5).

## Results

### Study Cohort

A total of 67 full-length endoscopic videos were collected from 59 patients with UC, comprising approximately 1.02 million video frames. Baseline demographic and clinical characteristics are summarised in Table 1.

The majority of procedures (89%) were performed for disease activity assessment, while 11% were undertaken for dysplasia surveillance. 53% were female, with a median age of 43 years (range 19–80). Mean haemoglobin was 133 g/L (range 80–161 g/L), mean C-reactive protein (CRP) was 13 mg/L (range <5–80 g/L), and mean faecal calprotectin was 1391 µg/g (range 15–6000 µg/g).

At the time of endoscopy, 5 patients were not receiving treatment for UC, 33 were receiving topical therapy, 20 biologic or small-molecule therapies, and 1 systemic corticosteroids alone.

### Frame-level quality control and endoscopic severity assessment

The scorable-frame model achieved an accuracy of 0.91 (95% CI 0.87–0.95), sensitivity of 0.90, specificity of 0.90 and AUROC of 0.90 (Supplementary Table 6).

For ordinal MES grading (0–3), the frame-level model achieved an accuracy of 0.89 (95% CI 0.83–0.94), weighted sensitivity of 0.89, weighted specificity of 0.96, weighted NPV of 0.96 and weighted PPV of 0.90. Agreement between AI predictions and expert assessment was almost perfect, with a quadratic weighted kappa (QWK) of 0.94 (95% CI 0.91–0.97) (Supplementary Table 7).

For UCEIS severity grading using four categories (0, 1–3, 4–5 and 6–8), the model achieved an accuracy of 0.83 (95% CI 0.74–0.92), weighted sensitivity of 0.83, weighted specificity of 0.94, weighted NPV of 0.93 and weighted PPV of 0.84. Agreement was similarly high, with a QWK of 0.92 (95% CI 0.87–0.96) (Supplementary Table 8).

### Continuous Disease Score (CDS)

Beyond categorical endoscopic assessment, the CDS framework enabled continuous quantification of inflammatory burden throughout the colon. Application of the trained model to full-length HD-WLE videos enabled continuous visualisation of disease activity across the entire examination. Predicted MES and UCEIS scores were displayed as a temporal scoring ribbon aligned with video playback, while non-scorable regions were simultaneously identified (Figure 2). This allowed disease severity and spatial distribution to be assessed continuously throughout the examination rather than at isolated time points or selected frames.

**Figure 2.**
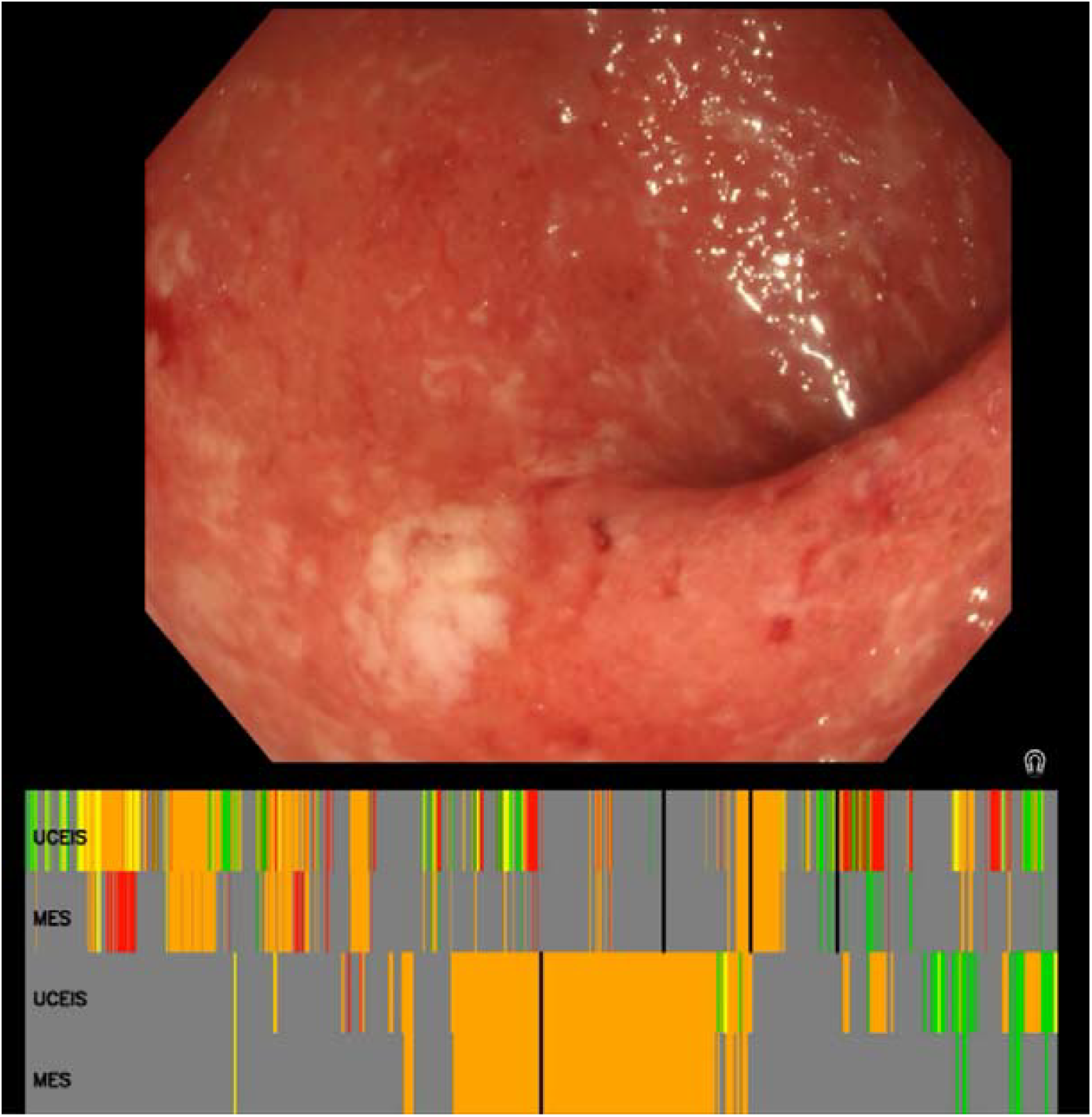
Video snapshot with continuous scoring ribbon. Predicted UCEIS and MES scores are shown over time for the full-length video (top) and a corresponding 20-second segment (bottom), with the snapshot location indicated by the vertical line.

Representative CDS outputs are shown in Figure 3. CDS profiles revealed marked differences in cumulative inflammatory burden and disease extent among patients with similar categorical endoscopic severity scores. In the examples shown, all patients met criteria for severe disease based on conventional ordinal scoring (MES 3 and UCEIS ≥6), yet demonstrated substantially different CDS values (86, 168 and 325), reflecting differences in both inflammatory extent and severity distribution throughout the colon. Whole-colon CDS profiles provided a summary of cumulative inflammatory burden across anatomical segments. Scatter plots of segment-level CDS values stratified by MES and UCEIS categories are shown in Supplementary Figures 1-2. CDS increased with worsening endoscopic severity for both scoring systems, but separation between severity categories was more distinct for UCEIS than for MES, with CDS values overlapping considerably across MES grades, particularly at the lower end of the range.

**Figure 3.**
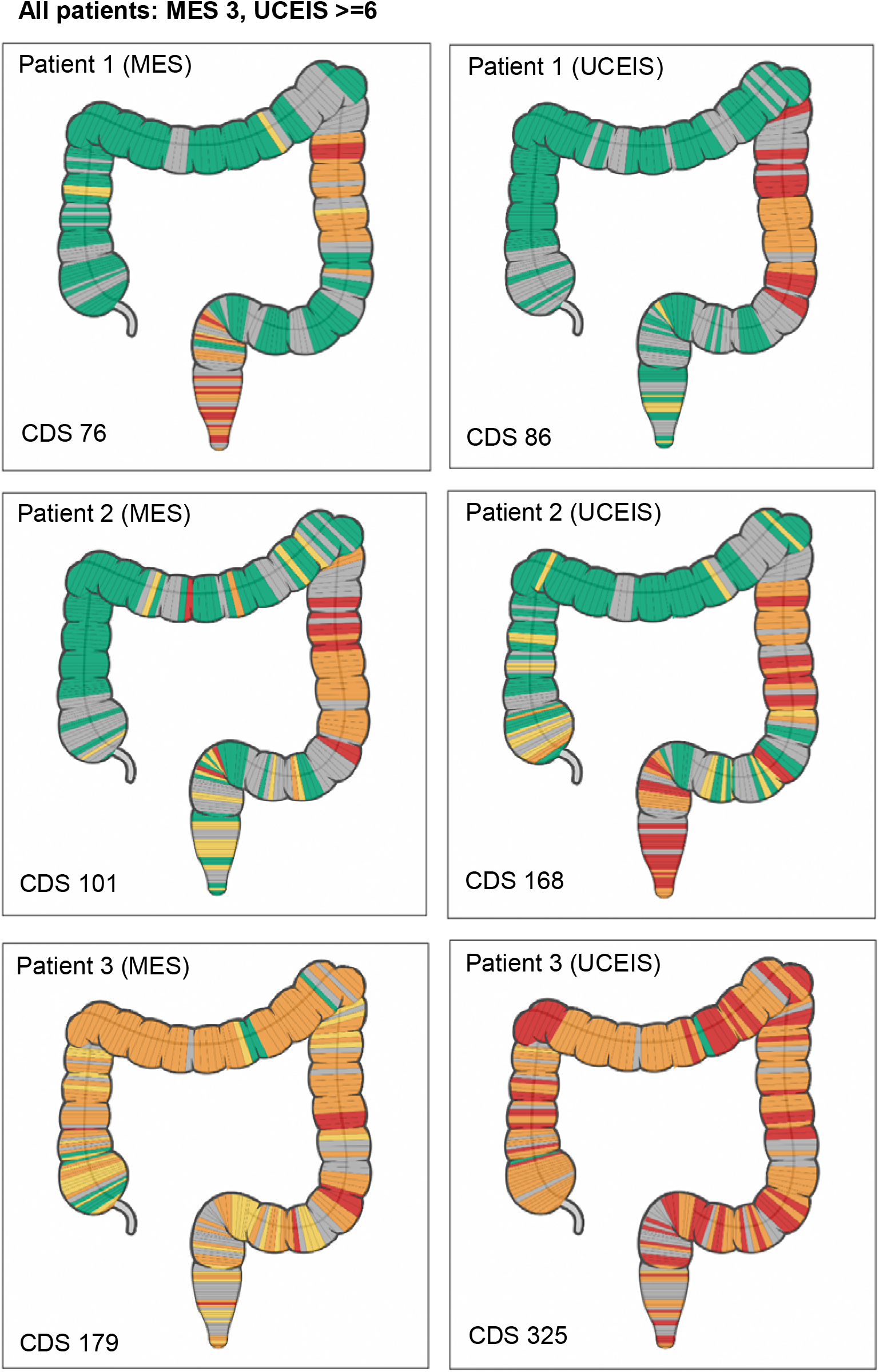
Whole-colon Continuous Disease Score (CDS) derived from UCEIS predictions in three patients. In all cases, categorical scores indicated severe disease (MES 3, UCEIS ≥6). CDS values differed substantially (86, 168, 325), reflecting differences in cumulative inflammatory burden and spatial extent of disease across the colon.

### Assessment of disease activity for anatomic segment videos

For anatomically defined video segments, the model detected endoscopic remission defined by MES=0 with an accuracy of 0.88 (95% CI 0.80–0.96), sensitivity of 0.88 (95% CI 0.78– 0.96), specificity of 0.87, NPV of 0.89, PPV of 0.89 and Cohen’s kappa of 0.76 (95% CI 0.56–0.92).

For remission defined by UCEIS≤1, accuracy was 0.92 (95% CI 0.84–0.98), sensitivity was 0.92 (95% CI 0.83–0.98), specificity was 0.92, NPV was 0.92, PPV was 0.93 and Cohen’s kappa was 0.84 (95% CI 0.67–0.96) (Table 2B).

**Table 2.** Performance of ML model for detection of endoscopic remission.

A. 20-second video clips
| Score | Accuracy (95% CI) | Sensitivity (95% CI) | Specificity | PPV | NPV | Cohen's $\kappa$ (95% CI) |
| --- | --- | --- | --- | --- | --- | --- |
| MES 0 | 0.89 (0.83–0.94) | 0.88 (0.82–0.94) | 0.88 | 0.90 | 0.90 | 0.78 (0.64–0.90) |
| UCEIS $\leq 1$ | 0.86 (0.79–0.92) | 0.85 (0.78–0.92) | 0.85 | 0.88 | 0.88 | 0.72 (0.56–0.85) |

B. Whole anatomical segments
| Score | Accuracy (95% CI) | Sensitivity (95% CI) | Specificity | PPV | NPV | Cohen's $\kappa$ (95% CI) |
| --- | --- | --- | --- | --- | --- | --- |
| MES 0 | 0.88 (0.80–0.96) | 0.88 (0.78–0.96) | 0.87 | 0.89 | 0.89 | 0.76 (0.56–0.92) |
| UCEIS $\leq 1$ | 0.92 (0.84–0.98) | 0.92 (0.83–0.98) | 0.92 | 0.93 | 0.92 | 0.84 (0.67–0.96) |

### Correspondence between AI-derived endoscopic activity and histological remission

To evaluate biological correspondence between AI-derived endoscopic activity and histological inflammation, biopsy-aligned video clips were analysed and compared with matched histopathology assessments.

For biopsy-aligned 20-second clips, the MES remission classifier achieved an accuracy of 0.89 (95% CI 0.83–0.94), sensitivity of 0.88 (95% CI 0.82–0.94), specificity of 0.88, PPV of 0.90, NPV of 0.90 and Cohen’s kappa of 0.78 (95% CI 0.64–0.90). The corresponding UCEIS remission classifier achieved an accuracy of 0.86 (95% CI 0.79–0.92), sensitivity of 0.85 (95% CI 0.78–0.92), specificity of 0.85, PPV of 0.88, NPV of 0.88 and Cohen’s kappa of 0.72 (95% CI 0.56–0.85) (Table 2A).

AI-derived endoscopic remission outputs were evaluated against each binary histological remission endpoint. For AI-derived MES remission, AUROC values were 0.84 (95% CI 0.78– 0.89), 0.84 (95% CI 0.79–0.89) and 0.87 (95% CI 0.82–0.91) for SGS, NHI and RHI, respectively. Corresponding AUROC values for AI-derived UCEIS remission, were 0.85 (95% CI 0.79–0.91), 0.84 (95% CI 0.78–0.90) and 0.87 (95% CI 0.82–0.92), respectively (Table 3).

**Table 3.**
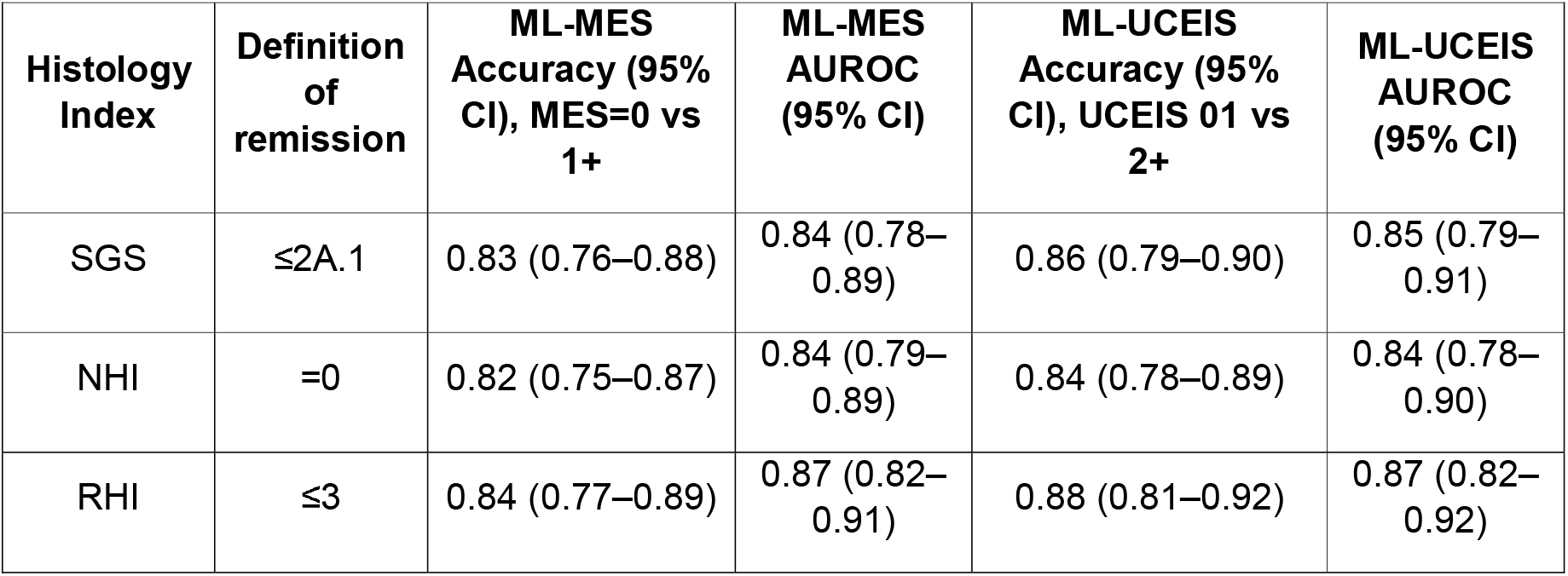
Correspondence between AI endoscopic scores and histological remission (senior histopathologist reference standard)

| Histology Index | Definition of remission | ML-MES Accuracy (95% CI), MES=0 vs 1+ | ML-MES AUROC (95% CI) | ML-UCEIS Accuracy (95% CI), UCEIS 01 vs 2+ | ML-UCEIS AUROC (95% CI) |
| --- | --- | --- | --- | --- | --- |
| SGS | $\leq 2A.1$ | 0.83 (0.76–0.88) | 0.84 (0.78–0.89) | 0.86 (0.79–0.90) | 0.85 (0.79–0.91) |
| NHI | =0 | 0.82 (0.75–0.87) | 0.84 (0.79–0.89) | 0.84 (0.78–0.89) | 0.84 (0.78–0.90) |
| RHI | $\leq 3$ | 0.84 (0.77–0.89) | 0.87 (0.82–0.91) | 0.88 (0.81–0.92) | 0.87 (0.82–0.92) |

## Discussion

This study developed and validated an artificial intelligence (AI) system for automated endoscopic assessment of disease activity in ulcerative colitis (UC) using full-length high-definition white-light endoscopy (HD-WLE) videos. The system achieved high accuracy for detecting endoscopic remission and for grading disease activity according to both the Mayo Endoscopic Subscore (MES) and the Ulcerative Colitis Endoscopic Index of Severity (UCEIS). The AI model also distinguished histological remission with strong agreement to expert histopathological assessment, demonstrating its potential to bridge the gap between endoscopic and histological evaluation. To our knowledge, this is among the first studies to apply a deep learning model to unaltered, full-length colonoscopy videos for both endoscopic and histological prediction across the entire colon in UC.

Previous studies have established the feasibility of AI-assisted evaluation of mucosal inflammation in UC but were largely limited to still images or short clips from selected colonic segments, while more recent models have extended these approaches using virtual chromoendoscopy or segment-level analysis. However, most of these systems required manual selection of representative images or depend on specialised imaging modalities that are not routinely utilised in standard clinical practice for IBD assessment. The present study advances the field by demonstrating that accurate, segment-by-segment and whole-colon evaluation can be achieved using routine HD-WLE alone, without image pre-selection or enhancement [7–11, 13].

Our findings compare favourably with previously reported AI models, achieving accuracies of 0.86–0.92 for endoscopic remission detection and κ values between 0.72 and 0.84, consistent with or exceeding interobserver agreement among expert endoscopists [19]. The continuous disease score (CDS) allows visualisation of disease distribution and severity gradients.

The CDS provides a clinically meaningful quantification of inflammatory burden that captures disease extent and heterogeneity not represented by categorical indices alone (Supplementary Figures 1-2) [5, 12]. A system performing continuous disease assessment may improve evaluation and prediction of outcomes in clinical practice and trials.

Representative cases shown in Figure 3 illustrate an important limitation of categorical endoscopic indices in full-length examinations. Although all three examples met criteria for severe disease based on ordinal UCEIS categorisation (total score ≥6), the CDS distinguished substantial differences in inflammatory extent and cumulative severity across the colon. This demonstrates how patients assigned to the same categorical severity class may nonetheless exhibit very different inflammatory phenotypes when disease distribution and exposure are taken into account. Clinically, CDS-based visualisation may better capture overall inflammatory burden and spatial heterogeneity than single-point ordinal scores. These examples illustrate the additional descriptive granularity provided by CDS rather than establish outcome-based thresholds or clinical decision cut-offs. Furthermore, the model’s strong performance in predicting histological remission underscores the potential for AI to reduce reliance on multiple mucosal biopsies where there is endoscopic remission, which are resource-intensive and impractical for real-time decision making [6, 9, 20].

Objective and reproducible quantification of mucosal healing is increasingly recognised as a critical treatment target in UC; however, widely used human scoring systems such as MES and UCEIS remain limited by subjectivity and interobserver variability. Central reading can mitigate these issues in clinical trials but is impractical for routine clinical use. Automated, AI-assisted scoring offers a practical solution by standardising interpretation and providing consistent outputs irrespective of operator experience. The ability to infer histological remission from endoscopic imaging represents an important step toward real-time biopsy-sparing evaluation, which could ultimately facilitate immediate therapeutic decisions at the point of care.

Several aspects strengthen this study. The dataset comprised unaltered, full-length HD-WLE videos captured in real-world clinical settings. The study design included independent, blinded scoring by experienced IBD endoscopists and histopathologists using multiple validated indices (MES, UCEIS, SGS, NHI, RHI), providing robust reference standards.

Model performance was evaluated using established classification metrics, and the study adhered to both CLAIM and TRIPOD guidelines, ensuring transparent reporting of AI development and validation.

Nevertheless, limitations must be acknowledged. Although the dataset included patients from two hospital sites, it was derived from a single healthcare network and may not encompass the full variability of UC presentations, endoscope types, or imaging conditions seen internationally. The sample size, while substantial for a video-based AI study, remains modest compared to large-scale imaging datasets, and external validation on independent multicentre cohorts is required. The system was evaluated retrospectively, and real-time performance in live endoscopy remains to be tested. Additionally, while histological differentiation was encouraging, the model did not incorporate clinical or biochemical parameters, which may further improve predictive accuracy when combined in a multimodal framework. Furthermore, although CDS characterises inflammatory phenotype differently, its clinical utility in predicting disease outcomes requires further validation.

Future work should focus on multicentre prospective validation and integration into central reading workstreams and clinical endoscopy practice. Future studies are needed to determine whether spatially resolved disease assessment may improve understanding of treatment response, support more personalised therapeutic strategies, and provide additional prognostic information beyond conventional categorical endoscopic indices.

Incorporation of multimodal data, including clinical symptoms, biomarkers, and histopathological feedback, may refine predictive performance and enable more comprehensive disease modelling. Longitudinal application of CDS could provide a more sensitive, quantitative biomarker for treatment response and relapse prediction. Ultimately, combining AI-derived endoscopic and histological predictions with clinical data could lead to a fully integrated precision-monitoring tool for ulcerative colitis.

In conclusion, this study demonstrates that a deep-learning based AI system can accurately assess endoscopic disease activity and predict histological remission from full-length HD-WLE colonoscopy videos. The model provides an objective, reproducible, and spatially resolved assessment of mucosal inflammation that aligns with expert evaluation. Integration of such AI systems into clinical practice and clinical trials could enhance the reliability of disease activity assessment, reduce interobserver variability, and enable real-time, biopsy-sparing evaluation of mucosal healing in ulcerative colitis.

## Data Availability Statement

The PAIR-IBD endoscopy system described in this study is the intellectual property of Perspectum Ltd and is the subject of a patent application (GB2608153.9). Summary data are included in the manuscript or uploaded as supplementary information (Supplementary Figures 1-2). Additional data underlying this study, including the WHITS (NCT05000242) video and annotation dataset, are proprietary and not publicly available, but may be considered for sharing on a case-by-case basis for legitimate research or commercial collaboration, subject to a data sharing agreement and applicable governance and ethical approvals (REC: 20/LO/0349). Requests should be directed to the corresponding author.

## Declaration of Generative AI Use

Generative AI (ChatGPT, OpenAI) was used to assist with language and grammar editing and the preparation of selected graphical elements. All AI-assisted content was reviewed and approved by the authors. Generative AI was not used for data analysis or scientific interpretation.

## References

1. Ng SC, Shi HY, Hamidi N et al. Worldwide incidence and prevalence of inflammatory bowel disease in the 21st century: a systematic review of population-based studies. The Lancet 2017; 390: 2769–2778

2. Turner D, Ricciuto A, Lewis A et al. STRIDE-II: an update on the Selecting Therapeutic Targets in Inflammatory Bowel Disease (STRIDE) Initiative of the International Organization for the Study of IBD (IOIBD): determining therapeutic goals for treat-to-target strategies in IBD. Gastroenterology 2021; 160: 1570–1583

3. Travis SP, Schnell D, Krzeski P et al. Developing an instrument to assess the endoscopic severity of ulcerative colitis: the Ulcerative Colitis Endoscopic Index of Severity (UCEIS). Gut 2012; 61: 535–542

4. Travis SP, Schnell D, Krzeski P et al. Reliability and initial validation of the ulcerative colitis endoscopic index of severity. Gastroenterology 2013; 145: 987–995

5. Lobatón T, Bessissow T, De Hertogh G et al. The Modified Mayo Endoscopic Score (MMES): a new index for the assessment of extension and severity of endoscopic activity in ulcerative colitis patients. Journal of Crohn’s and Colitis 2015; 9: 846–852

6. Bossuyt P, Nakase H, Vermeire S et al. Automatic, computer-aided determination of endoscopic and histological inflammation in patients with mild to moderate ulcerative colitis based on red density. Gut 2020; 69: 1778–1786

7. Stidham RW, Liu W, Bishu S et al. Performance of a deep learning model vs human reviewers in grading endoscopic disease severity of patients with ulcerative colitis. JAMA network open 2019; 2: e193963

8. Takenaka K, Ohtsuka K, Fujii T et al. Development and validation of a deep neural network for accurate evaluation of endoscopic images from patients with ulcerative colitis. Gastroenterology 2020; 158: 2150–2157

9. Iacucci M, Cannatelli R, Parigi TL et al. A virtual chromoendoscopy artificial intelligence system to detect endoscopic and histologic activity/remission and predict clinical outcomes in ulcerative colitis. Endoscopy 2023; 55: 332–341

10. Byrne MF, Panaccione R, East JE et al. Application of deep learning models to improve ulcerative colitis endoscopic disease activity scoring under multiple scoring systems. Journal of Crohn’s and Colitis 2023; 17: 463–471

11. Fan Y, Mu R, Xu H et al. Novel deep learning–based computer-aided diagnosis system for predicting inflammatory activity in ulcerative colitis. Gastrointestinal Endoscopy 2023; 97: 335–346

12. Stidham RW, Cai L, Cheng S et al. Using computer vision to improve endoscopic disease quantification in therapeutic clinical trials of ulcerative colitis. Gastroenterology 2024; 166: 155–167. e152

13. Takenaka K, Fujii T, Kawamoto A et al. Deep neural network for video colonoscopy of ulcerative colitis: a cross-sectional study. The lancet Gastroenterology & hepatology 2022; 7: 230–237

14. Maeda Y, Ditonno I, Puga-Tejada M et al. Artificial intelligence-enabled advanced endoscopic imaging to assess deep healing in inflammatory bowel disease. eGastroenterology 2024; 2: e100090. doi:10.1136/egastro-2024-100090

15. ClinicalTrials.gov. West Hertfordshire Inflammatory Bowel Disease Technology Study (WHITS). In

16. Windell D, Magness A, Li R et al. AI quantification of inflammatory and architectural features in ulcerative colitis distinguishes active disease from remission. medRxiv 2026. 2026.2001. 2027.26344949

17. Mongan J, Moy L, Kahn Jr CE. Checklist for artificial intelligence in medical imaging (CLAIM): a guide for authors and reviewers. In: Radiological Society of North America; 2020: e200029

18. Collins GS, Reitsma JB, Altman DG et al. Transparent reporting of a multivariable prediction model for individual prognosis or diagnosis (TRIPOD): the TRIPOD statement. Journal of British Surgery 2015; 102: 148–158

19. Hashash JG, Yu Ci Ng F, Farraye FA et al. Inter- and Intraobserver Variability on Endoscopic Scoring Systems in Crohn’s Disease and Ulcerative Colitis: A Systematic Review and Meta-Analysis. Inflamm Bowel Dis 2024; 30: 2217–2226. doi:10.1093/ibd/izae051

20. Puga-Tejada M, Majumder S, Maeda Y et al. Artificial intelligence–enabled histology exhibits comparable accuracy to pathologists in assessing histological remission in ulcerative colitis: a systematic review, meta-analysis, and meta-regression. Journal of Crohn’s and Colitis 2025; 19: jjae198

