## Supplementary Information for "Deep learning-based assessment of ulcerative colitis activity from full-length endoscopic videos with spatial characterisation and histological correlation"

Supplementary Table 1. Frame labels distribution

| **Dataset** | **MES 0** | **MES 1** | **MES 2** | **MES 3** | **UCEIS 0** | **UCEIS 1-3** | **UCEIS 4-5** | **UCEIS 6-8** |
| --- | --- | --- | --- | --- | --- | --- | --- | --- |
| Train | 306 | 111 | 76 | 34 | 294 | 98 | 112 | 21 |
| Validation | 105 | 40 | 26 | 10 | 97 | 32 | 39 | 5 |
| Test (held out) | 101 | 37 | 25 | 9 | 96 | 32 | 38 | 7 |

Supplementary Table 2. Anatomical segment videos labels distribution

| **Dataset** | **MES 0** | **MES >=1** | **UCEIS 01** | **UCEIS >1** |
| --- | --- | --- | --- | --- |
| Train | 39 | 44 | 36 | 45 |
| Validation | 9 | 9 | 9 | 11 |
| Test (held out) | 24 | 26 | 22 | 28 |

Supplementary Table 3. 20-sec video clip labels distribution

| **Dataset** | **MES 0** | **MES >=1** | **UCEIS 01** | **UCEIS >1** |
| --- | --- | --- | --- | --- |
| Train | 74 | 86 | 84 | 73 |
| Validation | 26 | 30 | 31 | 27 |
| Test (held out) | 46 | 54 | 54 | 47 |

### Supplementary Table 4. CLAIM checklist for Artificial Intelligence in Medical Imaging

| **CLAIM Item** | **Description** | **Addressed (Yes/No)** | **Location in Manuscript** |
| --- | --- | --- | --- |
| Study design | Retrospective observational study | Yes | Methods |
| Data source | Real-world clinical endoscopy videos | Yes | Methods |
| Ground truth | Expert endoscopist and histopathologist scoring | Yes | Methods |
| Dataset partitioning | Independent training, validation, test sets | Yes | Methods |
| Model description | CNN-based multi-stage architecture | Yes | Methods, Fig. 1 |
| Evaluation metrics | Accuracy, AUROC, sensitivity, specificity, kappa | Yes | Statistical Analysis |
| Bias and limitations | Single-network dataset, retrospective design | Yes | Discussion |
| Transparency | Model outputs and failure modes described | Yes | Results |

Supplementary Table 5. TRIPOD checklist for prediction model development and validation

| **TRIPOD Item** | **Description** | **Addressed (Yes/No)** | **Location** |
| --- | --- | --- | --- |
| Objective definition | Endoscopic and histologic prediction | Yes | Objectives |
| Participants | UC patients undergoing endoscopy | Yes | Methods |
| Predictors | HD-WLE video features | Yes | Methods |
| Outcome definition | MES, UCEIS, SGS, NHI, RHI | Yes | Methods |
| Sample size | Video- and segment-level analyses | Yes | Results |
| Model development | CNN-based supervised learning | Yes | Methods |
| Validation | Independent test set | Yes | Results |
| Performance reporting | AUROC, kappa, CI | Yes | Results |

Supplementary Table 6. Quality control model performance (scorable vs non-scorable):

| **Accuracy (95% CI)** | **Sensitivity** | **Specificity** | **AUROC** |
| --- | --- | --- | --- |
| 0.91 (0.87–0.95) | 0.90 | 0.90 | 0.90 |

#### Supplementary Table 7. Performance of ML model for ordinal MES grading (0–3), frame level.

| **Metric** | **Value (95% CI)** |
| --- | --- |
| Accuracy | **0.89** (0.83–0.94) |
| AUROC (macro) | **0.86** (0.80–0.93) |
| Cohen’s kappa | **0.81** (0.71–0.90) |
| Quadratic weighted kappa | **0.94** (0.91–0.97) |
| Weighted sensitivity (TPR) | **0.89** |
| Weighted specificity (TNR) | **0.96** |
| Weighted PPV | **0.90** |
| Weighted NPV | **0.96** |

Supplementary Table 8. Performance of ML model for categorised UCEIS severity, frame level.

| **Metric** | **Value (95% CI)** |
| --- | --- |
| Accuracy | **0.83** (0.74–0.92) |
| AUROC (macro) | **0.89** (0.83–0.94) |
| Cohen’s kappa | **0.76** (0.63–0.86) |
| Quadratic weighted kappa | **0.92** (0.87–0.96) |
| Weighted sensitivity (TPR) | **0.83** |
| Weighted specificity (TNR) | **0.94** |
| Weighted PPV | **0.84** |
| Weighted NPV | **0.93** |

*UCEIS categories defined as: 0 (healthy/remission), 1–3 (mild), 4–5 (moderate), ≥6 (severe).*

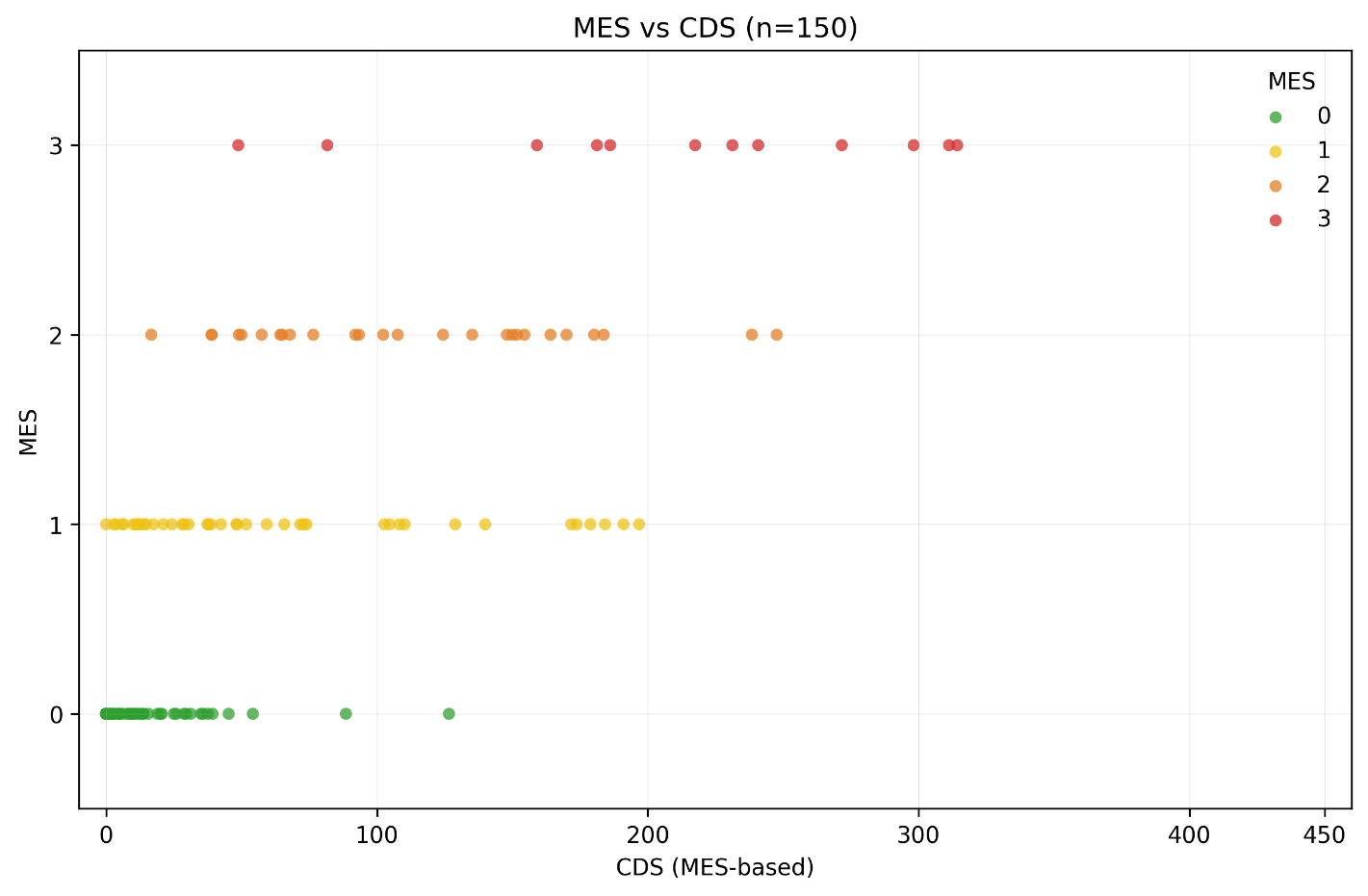

Supplementary Figure 1. Relationship between MES-based CDS and MES at segment level.

Scatter plot showing the relationship between CDS derived from MES predictions and adjudicated MES grades (0–3) for individual anatomical segments. Point colours indicate MES categories (0 = green, 1 = yellow, 2 = orange, 3 = red).

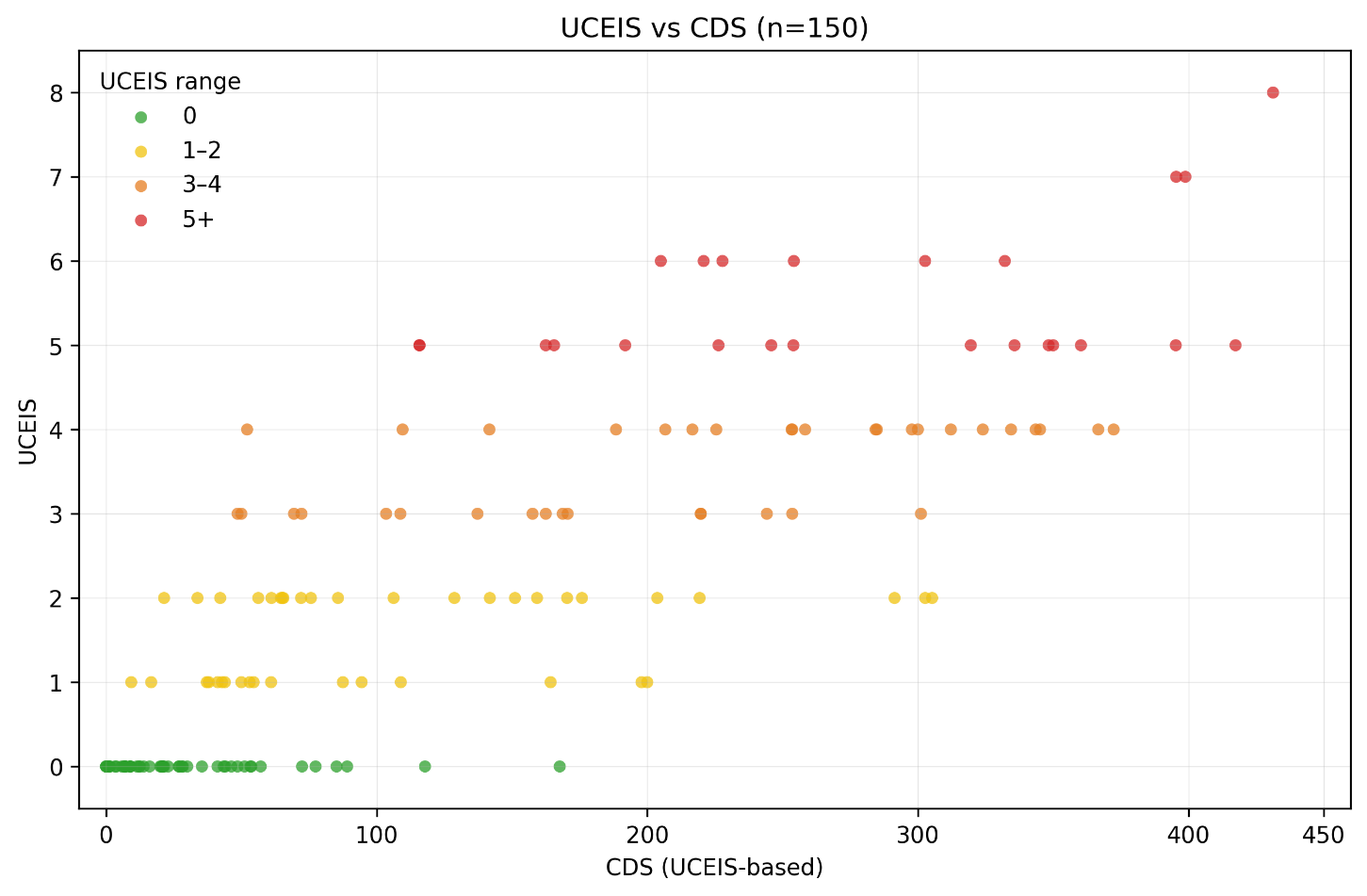

Supplementary Figure 2. Relationship between UCEIS-based CDS and adjudicated UCEIS score at segment level.

Scatter plot showing the relationship between CDS derived from UCEIS predictions and adjudicated UCEIS scores (0–8) for individual anatomical segments. Point colours indicate UCEIS severity categories (0 = green, 1–2 = yellow, 3–4 = orange, ≥5 = red).
